# Medication Guideline Adherence Among Patients with Markedly Elevated Blood Pressure in A Real-World Setting

**DOI:** 10.1101/2022.02.16.22271094

**Authors:** Yuan Lu, Chenxi Huang, Yuntian Liu, César Caraballo, Shiwani Mahajan, Daisy Massey, Erica S. Spatz, Oyere Onuma, Joseph S. Ross, Wade L. Schulz, Harlan M. Krumholz

## Abstract

**Objective:** To evaluate medication guideline adherence among ambulatory patients with markedly elevated blood pressure (BP), overall and by patient characteristics.

**Design:** Population-based, retrospective cohort study.

**Setting:** Yale New Haven Health System.

**Participants:** Adult patients aged 18-85 years with markedly elevated BP (defined as two consecutive outpatient visits with systolic BP ≥160 mmHg or diastolic BP ≥100 mmHg) between October 1^st^, 2015 and December 31^st^, 2018.

**Main outcome measures:** We assessed the number and class of antihypertensive drugs (previously taken and newly prescribed) prior to 90 days of the second visit with BP ≥160/100 mmHg. Among patients treated with two-drug class regimens, we assessed the proportion of patients on guideline-recommended two drug classes, overall and stratified by documentation of prior myocardial infarction (MI), diabetes, chronic kidney disease (CKD), and uncomplicated hypertension (i.e., without MI, coronary artery disease, diabetes, CKD, and cerebrovascular disease).

**Results:** We identified 16,377 patients with markedly elevated BP. They had a mean age of 65.8 (SD: 14.5) years; 54.0% were female; and 69.4%, 19.6%, and 9.6% were White, Black, and Hispanic, respectively. Prior to 90 days of the second visit, 29.8% had no active antihypertensive drug prescription, 20.0% had one drug class prescribed, and 50.2% had two or more drug classes prescribed. Among patients prescribed one antihypertensive drug class, the most common drug class was angiotensin converting enzyme inhibitor (ACEI) or angiotensin receptor blocker (ARB), followed by calcium channel blocker (CCB). Among patients prescribed two antihypertensive drug classes, the most common treatment combinations were ACEI or ARB and thiazide diuretic (21.0%), followed by ACEI or ARB and CCB (20.6%). Guideline-recommended two-drug class combination therapy were prescribed in 54.3% of the treated population, with the highest proportion of 67.0% in patients with a prior MI and the lowest proportion of 48.2% among patients with CKD. Older age, lower body mass index, and lower BP were associated with lower prescription of guideline-recommended combination therapy.

**Conclusion:** Only half of patients with markedly elevated BP were prescribed guideline-recommended antihypertensive combination drugs. Major opportunities exist for improving the guideline adherence of antihypertensive drug prescription in this population.

**What this Paper Adds:** *Section 1: What is already known on this subject?:* Hypertension affects nearly one in two adults in the United States, of whom 20% have markedly elevated blood pressure (BP; defined as BP ≥160/100 mmHg). People with markedly elevated BP have increased risks of adverse cardiovascular and kidney events. Clinical guidelines recommend prescribing combination therapy with two or more antihypertensive agents for patients with markedly elevated BP.

*Section 2: What this study adds:* This study quantified the real-world medication guideline adherence among ambulatory patients with markedly elevated BP, using electronic health record data from a large health system in the United States. Prior to 90 days of the second visit with BP ≥160/100 mmHg, nearly 30% of patients had no active antihypertensive drug prescription. Among patients prescribed at least one antihypertensive drug class, guideline-recommended two-drug class combination therapy were prescribed in 54% of patients, with the highest proportion of 67% in patients with a prior myocardial infarction and the lowest proportion of 48% among patients with chronic kidney disease. These findings highlight a large missed opportunity for improving the guideline adherence of antihypertensive drug prescription in this population.

## BACKGROUND

Hypertension affects nearly one in two adults in the United States, of whom 20% have markedly elevated blood pressure (BP), defined as systolic BP ≥160 mmHg or diastolic BP ≥100 mm Hg.^1^ People with markedly elevated BP have increased risks of heart failure, stroke, atherosclerotic coronary artery disease, renal failure, and death compared to those with moderately elevated or normal BP.^2^ Therefore, it is a public health priority to target these high-risk people and ensure that they receive proper treatment to achieve BP control.

Current hypertension practice guidelines recommend combination therapy with two or more antihypertensive agents as the initial treatment for people with markedly elevated BP.^3 4^ The guidelines also recommend specific antihypertensive drug classes for patients based on their comorbidities and drug combinations with complementary mechanisms of action to achieve greater BP-lowering efficacy. For example, the guidelines recommend dual-combination therapy that combines an angiotensin converting enzyme inhibitor (ACEI) or angiotensin receptor blocker (ARB) with a calcium channel blocker (CCB) or a thiazide diuretic for patients with chronic kidney disease (CKD). While these guidelines are well-established, physicians’ guideline adherence is suboptimal, creating a critical barrier to patients achieving BP control goals.^5^

There is a lack of high-quality, real-world contemporary evidence assessing physician adherence to medication guidelines among people with markedly elevated BP. Studies have been limited by focusing on all people with hypertension, not systematically assessing medication intensity (number and class), or not assessing heterogeneity in different patient subgroups.^6-9^ Moreover, determinants of medication guideline adherence among people with markedly elevated BP may be multifactorial and have not been well characterized.

The wide adoption of electronic health records (EHR) provides an opportunity to evaluate medication prescription among patients with markedly elevated BP in real-world practice. Using EHR data from a large health system, we assessed real-world physician adherence to medication guidelines among patients with markedly elevated BP and identify factors associated with adherence, emphasizing patient characteristics and comorbid medical conditions.

## METHODS

### Data Source

We performed retrospective analyses of de-identified data on an EHR-based cohort of adult patients in the Yale-New Haven Health System (YNHHS). YNHHS is Connecticut’s largest healthcare system, consisting of five hospitals and one outpatient provider network. YNHHS has about 3 million outpatient visits from 100,000 unique patients annually. All YNHHS hospitals used a secure, centralized EHR system designed by the Epic Corporation to collect and store clinical and administrative claims data. The EHR data were transformed into the PCORnet common data model via extract/transform/load software.^10 11^ We used a versioned extract of the PCORnet data from October 1, 2015, when International Classification of Diseases-10th Edition-Clinical Modification (ICD-10-CM) diagnosis was introduced, through December 31, 2018.

### Cohort Definition

The study population included patients aged 18-85 years with at least two outpatient visits in the YNHHS between October 1^st^, 2015, and December 31^st^, 2018. We constructed a cohort of patients with markedly elevated BP, defined as patients who had two consecutive outpatient visits with systolic BP ≥160 mmHg or diastolic BP ≥100 mmHg during the study period.

Cohort entry (index date) for each patient was their second consecutive visit with BP ≥160/100 mmHg. Inclusion criteria for patients based on the index date included: 1) at least seven days but no more than six months between the two consecutive visits with systolic BP ≥160 mmHg or diastolic BP ≥100 mmHg; and 2) at least three months follow-up time after the index date (**Appendix Figure 1**).

We excluded adults who were pregnant or on dialysis during the study period. We also excluded inpatient, emergency department, or ambulatory surgery center BP values to reduce the risk of transiently elevated BP from acute medical conditions.^12 13^ We extracted patient demographics, past medical histories, vital signs, outpatient medications, and laboratory results from the EHR of all outpatient visits in which a patient met our criteria. If a patient had more than two BP measurements from a single visit, we used the mean of all measurements as the BP for that visit.

### Variable Definitions

#### Antihypertensive Medication Classification

Medication prescription orders (name, generic name, quantity, frequency, RxNorm) for each outpatient visit were extracted from the PCORnet common data model. The Yale Epic team routinely performed internal validation supplemented by manual audits to ensure the quality of medication data. To properly classify EHR-based medication prescription data into antihypertensive therapeutic indication and antihypertensive drug classes, we used a previously developed antihypertensive drug classification system based on RxNorm Concept Unique Identifier (RxCUI).^14^ We included only oral formulations to focus on outpatient medications, with the exception of transdermal clonidine patches, as some medications have ophthalmic dosage forms that are not indicated for hypertension.

We then applied the classification system to EHR-based medication prescription order data from patients in YNHHS and classified them into different drug classes, including ACEI, ARB, CCB, thiazide or thiazide-like diuretic (TD), beta-blocker (BB), alpha-1 blocker, and antiadrenergic agent. For combination drugs, we classified them into the multiple classes of the combination drugs.

We evaluated medication prescription of eligible patients. We defined patients receiving active prescription for antihypertensive drugs using two approaches. In approach 1, active prescription was defined when the prescription order start date was between 360 days prior to the index date and 90 days after the index date (**Appendix Figure 2**). In approach 2, active prescription was defined when the prescription order start date was between 90 days prior to the index date and 90 days after the index date. Compared with approach 2, approach 1 allowed us to include long-term prescription orders that were made more than 90 days before the index date (e.g., prescription order made 6 months before the index date and still active). We evaluated the performance of diferent approaches in a manual chart review of a random sample of 50 patients. Using clinicians’ manual judgements as the reference, the use of approach 1 to identify patients treated with antihypertensive medications had better performance characteristics compared with approach 2 (**Appendix Figure 3**). Hence, we reported the results of approach 1 in the main analysis, and the results of approach 2 in the sensitivity analysis.

#### Comorbidities

Data on patient comorbidities were assessed by identifying ICD-10 codes either listed as a billing diagnosis or on the patient problem list at any point in the year prior to the index visit for the following conditions: heart failure, diabetes mellitus, dyslipidemia, myocardial infarction (MI), coronary artery disease, cerebrovascular disease, atrial fibrillation, CKD, chronic obstructive pulmonary disease, peripheral arterial disease, angina, hemorrhagic stroke, ischemic stroke, depression, dementia, hypertensive retinopathy, and substance use disorder (see detail definitions in **Appendix Table 1**).

#### DefinitionofPreferredCombinationTherapybyComorbidity

We defined the preferred antihypertensive combination therapy for patients with history of MI, diabetes, CKD, and uncomplicated hypertension based on the 2017 American College of Cardiology /American Heart Association (ACC/AHA) hypertension guidelines.^3^ History of MI, diabetes, and CKD was defined as diagnosis of MI, diabetes, and CKD at any point prior to the index visit. Uncomplicated hypertension was defined as lack of MI, coronary artery disease, diabetes, CKD, and cerebrovascular disease. We considered ACEI/ARB and CCB/TD as the preferred two drug classes for patients with diabetes and CKD. Although the 2017 ACC/AHA hypertension guidelines emphasized ACEI/ARB for patients with diabetes with proteinuria,^3^ we conducted the analyses among all people with diabetes regardless of proteinuria because we considered that our definition of preferred two drug classes did not differ by proteinuria status. Among patient with prior MI, we considered ACEI/ARB and BB as the preferred two drug classes regardless of time since MI, consistent with the most recent MI and hypertension treatment guidelines in the US.^3 15^ We considered ACEI/ARB and CCB/TD, or BB and CCB/TD as the preferred two drug classes for patients with uncomplicated hypertension (**Appendix Table 2**).

### Statistical Analysis

We first described the sociodemographic and clinical characteristics, number and class of antihypertensive drugs prescribed among patients with markedly elevated BP. Among patients treated with two drug class regimens, we assessed the proportion of patients on guideline preferred vs. non-preferred combination therapy by history of MI, diabetes, CKD, and uncomplicated hypertension. We calculated number and percentage of patients whose medications are concordant with guidelines and stratified the analysis by patients’ demographic characteristics. Finally, we fitted a gradient boosted machine model to identify patient characteristics associated with prescription of guideline-recommended combination therapy. A gradient boosted machine model was chosen because this tree-based machine learning algorithm was flexible and efficient in learning interactions between different features in the EHR data. We considered a binary variable indicating whether a patient was prescribed a guideline-recommended combination therapy as the outcome variable and patient characteristics such as age, sex, race, ethnicity, insurance type, preferred language, body mass index (BMI) category, smoking status, and comorbidities as candidate features in the model. We evaluated the relative importance of patient characteristics in predicting prescription of guideline-recommended combination therapy by calculating the permutation feature importance scores. We showed the relationships between patient characteristics and outcome using partial dependence plots. All analyses were performed using R 3.6.0. This study received an exemption for review from the Institutional Review Board at Yale University because EHR data are de-identified. The study was reported following the STROBE (Strengthening the Reporting of Observational Studies in Epidemiology) reporting guidelines.^16^

## RESULTS

### Population Characteristics

We identified 16,377 patients who met the study criteria from 2015-2018 and were included in the final analysis (**Appendix Figure 4**). The mean age at the index date was 65.8 (SD, 14.5) years; 54.0% were female; 69.4%, 19.6%, and 9.6% were White, Black, and Hispanic, respectively. Overall, a total of 44.8% patients had a BMI of ≥30.0 kg/m^2^; 41.8% had dyslipidemia, 28.0% had diabetes, 11.1% had CKD, and 2.3% had experienced a prior MI. Mean (SD) systolic and diastolic BP of the population at the index date was 166.8 (13.1) mmHg and 88.6 (13.3) mmHg, respectively. The characteristics of patients with history of MI, diabetes, CKD, or uncomplicated hypertension are described in **Table 1**.

**Table 1.** Characteristics of patients with markedly elevated blood pressure.

| Characteristics, N (%) | Overall<br>(N=16,377) | Prior MI<br>(N=372) | Diabetes<br>(N=4,577) | CKD<br>(N=1,815) | Uncomplicated<br>hypertension*<br>(N=9,898) |
| --- | --- | --- | --- | --- | --- |
| Age, yrs., mean (SD) | 65.8 (14.5) | 70.4 (13.0) | 66.7 (13.0) | 72.1 (13.1) | 64.0 (15.0) |
| **Age group, N (%)** |  |  |  |  |  |
| 18-44 years | 1,312 (8.0) | 8 (2.2) | 224 (4.9) | 53 (2.9) | 1,060 (10.7) |
| 45-64 years | 6,021 (36.8) | 111 (29.8) | 1,705 (37.3) | 452 (24.9) | 3,891 (39.3) |
| >=65 years | 9,044 (55.2) | 253 (68.0) | 2,648 (57.9) | 1,310 (72.2) | 4,947 (50.0) |
| **Sex, N (%)** |  |  |  |  |  |
| Female | 8,849 (54.0) | 197 (53.0) | 2,368 (51.7) | 895 (49.3) | 5,618 (56.8) |
| Male | 7,528 (46.0) | 175 (47.0) | 2,209 (48.3) | 920 (50.7) | 4,280 (43.2) |
| **Race, N (%)** |  |  |  |  |  |
| Black | 3,204 (19.6) | 70 (18.8) | 1,272 (27.8) | 474 (26.1) | 1,695 (17.1) |
| White | 11,367 (69.4) | 256 (68.8) | 2,602 (56.8) | 1,140 (62.8) | 7,240 (73.1) |
| Others | 1,637 (10.0) | 44 (11.8) | 675 (14.7) | 192 (10.6) | 837 (8.5) |
| Unknown | 169 (1.0) | 2 (0.5) | 28 (0.6) | 9 (0.5) | 126 (1.3) |
| **Ethnicity, N (%)** |  |  |  |  |  |
| Hispanic | 1,566 (9.6) | 48 (12.9) | 666 (14.6) | 184 (10.1) | 772 (7.8) |
| Non-Hispanic | 14,486 (88.5) | 320 (86.0) | 3,856 (84.2) | 1,619 (89.2) | 8,880 (89.7) |
| Other/Unknown | 325 (2.0) | 4 (1.1) | 55 (1.2) | 12 (0.7) | 246 (2.5) |
| **Insurance type, N (%)** |  |  |  |  |  |
| Public (Medicare or Medicaid) | 10,778 (65.8) | 289 (77.7) | 3,269 (71.4) | 1,469 (80.9) | 5,988 (60.5) |
| Private | 5,085 (31.0) | 74 (19.9) | 1,146 (25.0) | 290 (16.0) | 3,600 (36.4) |
| Military | 55 (0.3) | 0 (0.0) | 11 (0.2) | 1 (0.1) | 39 (0.4) |
| None | 266 (1.6) | 7 (1.9) | 93 (2.0) | 22 (1.2) | 157 (1.6) |
| Others/Unknown | 193 (1.2) | 2 (0.5) | 58 (1.3) | 33 (1.8) | 114 (1.2) |
| **Preferred language, N (%)** |  |  |  |  |  |
| English | 15,203 (92.8) | 333 (89.5) | 4,052 (88.5) | 1,654 (91.1) | 9,359 (94.6) |
| Spanish | 779 (4.8) | 26 (7.0) | 389 (8.5) | 114 (6.3) | 323 (3.3) |
| Others | 350 (2.1) | 11 (3.0) | 125 (2.7) | 42 (2.3) | 189 (1.9) |
| Unknown | 45 (0.3) | 2 (0.5) | 11 (0.2) | 5 (0.3) | 27 (0.3) |
| **BMI category, N (%)** |  |  |  |  |  |
| ≥ 30 kg/m<sup>2</sup> | 7,342 (44.8) | 151 (40.6) | 2,641 (57.7) | 814 (44.8) | 4,064 (41.1) |
| 25-<30 kg/m<sup>2</sup> | 5,038 (30.8) | 120 (32.3) | 1,232 (26.9) | 542 (29.9) | 3,145 (31.8) |
| < 25 kg/m<sup>2</sup> | 3,328 (20.3) | 96 (25.8) | 565 (12.3) | 409 (22.5) | 2,200 (22.2) |
| Unknown | 669 (4.1) | 5 (1.3) | 139 (3.0) | 50 (2.8) | 489 (4.9) |
| **Smoking status, N (%)** |  |  |  |  |  |
| Current smoker | 488 (3.0) | 50 (13.4) | 168 (3.7) | 91 (5.0) | 220 (2.2) |
| Former smoker | 2,222 (13.6) | 145 (39.0) | 907 (19.8) | 495 (27.3) | 895 (9.0) |
| Never smoker | 299 (1.8) | 25 (6.7) | 122 (2.7) | 67 (3.7) | 99 (1.0) |
| Unknown | 13,368 (81.6) | 152 (40.9) | 3,380 (73.8) | 1,162 (64.0) | 8,684 (87.7) |
| **Comorbidities, N (%)** |  |  |  |  |  |
| Heart failure | 1,493 (9.1) | 171 (46.0) | 696 (15.2) | 483 (26.6) | 390 (3.9) |
| Diabetes mellitus | 4,577 (28.0) | 164 (44.1) | 4,577 (100.0) | 960 (52.9) | 0 (0.0) |
| Dyslipidemia | 6,852 (41.8) | 279 (75.0) | 2,901 (63.4) | 1,154 (63.6) | 2,758 (27.9) |
| Acute MI | 372 (2.3) | 372 (100.0) | 164 (3.6) | 92 (5.1) | 0 (0.0) |
| Coronary artery disease | 2,653 (16.2) | 293 (78.8) | 1,116 (24.4) | 585 (32.2) | 0 (0.0) |
| Cerebrovascular disease | 276 (1.7) | 22 (5.9) | 123 (2.7) | 66 (3.6) | 0 (0.0) |
| Atrial fibrillation | 1,633 (10.0) | 98 (26.3) | 534 (11.7) | 330 (18.2) | 651 (6.6) |
| Chronic kidney disease | 1,815 (11.1) | 92 (24.7) | 960 (21.0) | 1,815 (100.0) | 572 (5.8) |
| Chronic obstructive pulmonary disease | 1,236 (7.5) | 55 (14.8) | 467 (10.2) | 235 (12.9) | 507 (5.1) |
| Peripheral arterial disease | 1,015 (6.2) | 63 (16.9) | 454 (9.9) | 235 (12.9) | 302 (3.1) |
| Angina | 284 (1.7) | 50 (13.4) | 136 (3.0) | 57 (3.1) | 51 (0.5) |
| Hemorrhagic stroke | 91 (0.6) | 5 (1.3) | 30 (0.7) | 17 (0.9) | 0 (0.0) |
| Ischemic stroke | 606 (3.7) | 42 (11.3) | 262 (5.7) | 131 (7.2) | 0 (0.0) |
| Depression | 1,918 (11.7) | 64 (17.2) | 694 (15.2) | 283 (15.6) | 993 (10.0) |
| Dementia | 413 (2.5) | 22 (5.9) | 130 (2.8) | 82 (4.5) | 203 (2.1) |
| Hypertensive retinopathy | 71 (0.4) | 5 (1.3) | 46 (1.0) | 20 (1.1) | 13 (0.1) |
| Substance use disorder | 2,346 (14.3) | 94 (25.3) | 715 (15.6) | 292 (16.1) | 1,289 (13.0) |

Abbreviations: AMI, myocardial infarction; CKD, chronic kidney disease.
\* Uncomplicated hypertension was defined as lack of MI, coronary artery disease, diabetes, CKD, and cerebrovascular disease. The four comorbidity groups are not mutually exclusive. It is possible that a patient has more than one condition.

### Number and Class of Antihypertensive Medications Prescribed

Among women, 33.3% of those aged 18-44 years, 28.7% of those aged 45-64 years, and 29.5% of those aged 65 years or older were not prescribed any antihypertensive drugs prior to 90 days of the second visit (**Figure 1**). Among women who were prescribed antihypertensive drugs, 28.6% were prescribed one drug class, 27.8% were prescribed drugs from two drug classes, and 43.7% were prescribed three or more drug classes. Similarly, among men, 31.0% of those aged 18-44 years, 28.3% of those aged 45-64 years, and 31.8% of those aged 65 years or older were not prescribed any antihypertensive drugs prior to 90 days of the second visit. Among men who were prescribed antihypertensive drugs, 28.5% were prescribed one drug, 27.9% were prescribed drugs from two drug classes, and 43.6% were prescribed three or more drug classes.

**Figure 1.**
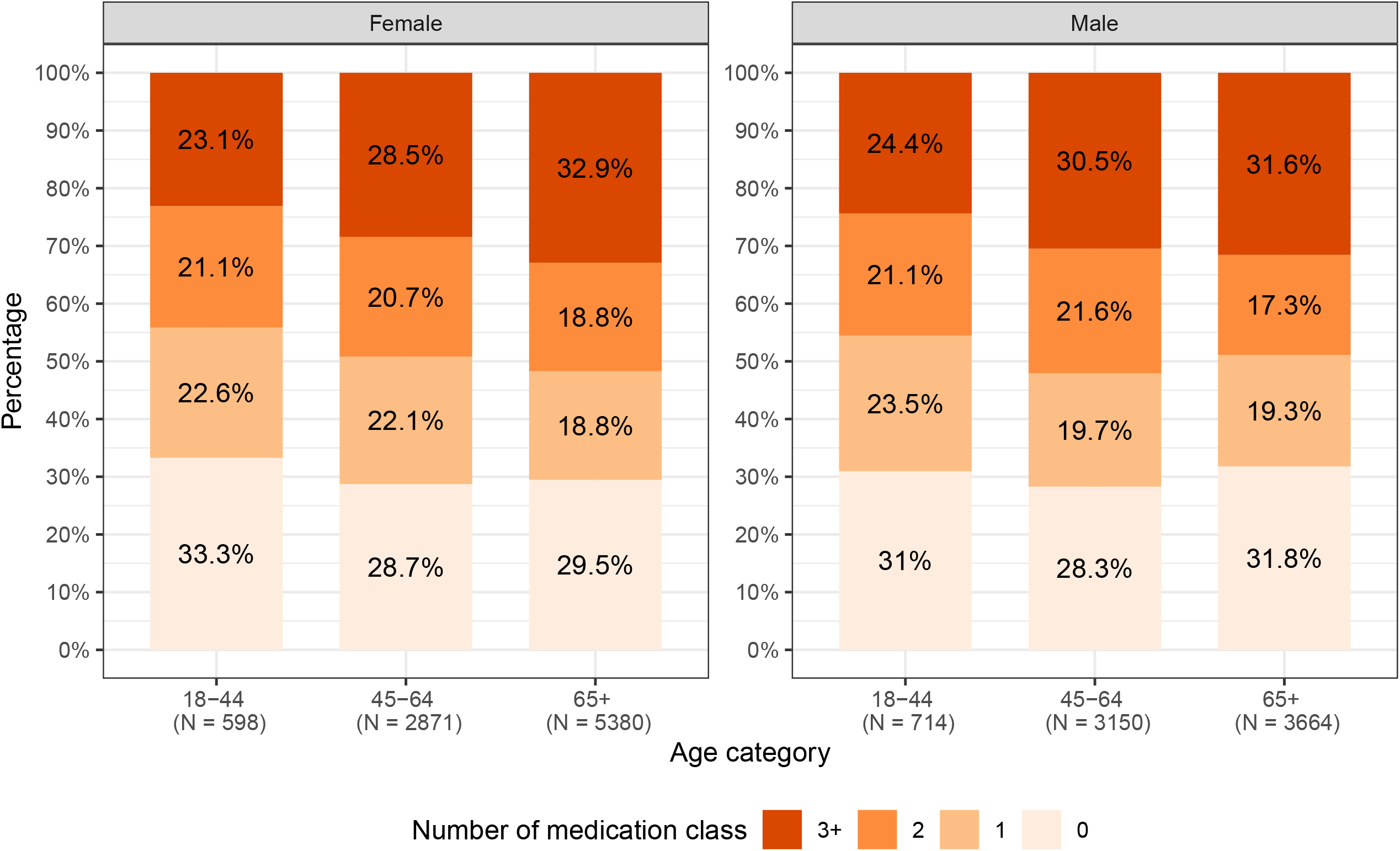
Number of antihypertensive medication class prescribed among patients with markedly elevated blood pressure.

Overall, 45.0% of patients with markedly elevated BP were prescribed an ACEI or ARB, 35.8% were prescribed a CCB, 36.7% were prescribed a BB, 24.4% were prescribed a TD, and 24.7% were prescribed other antihypertensive drugs (**Table 2**). Among patients who had a prior MI, the two most prescribed antihypertensive drug classes were beta-blocker (80.1%) and ACEI or ARB (72.0%), ACEI or ARB (58.3%) and beta-blocker (46.6%) among patients who had diabetes, beta-blocker (56.0%) and ACEI or ARB (37.1%) among patients who had CKD, and ACEI or ARB (37.0%) and CCB (30.0%) among those with uncomplicated hypertension.

**Table 2.** Antihypertensive medication classes prescribed among patients with markedly elevated blood pressure.

| Medication class | Overall<br>(N=16,377) | Prior MI<br>(N=372) | Diabetes<br>(N=4,577) | CKD<br>(N=1,815) | Uncomplicated<br>hypertension*<br>(N=9,898) |
| --- | --- | --- | --- | --- | --- |
| ACEI | 4,180 (25.5) | 166 (44.6) | 1,568 (34.3) | 511 (19.2) | 2,047 (20.7) |
| ARB | 3,629 (22.2) | 129 (34.7) | 1,262 (27.6) | 514 (18.5) | 1,825 (18.4) |
| ACEI or ARB | 7,369 (45.0) | 268 (72.0) | 2,670 (58.3) | 968 (37.1) | 3,663 (37.0) |
| CCB | 5,868 (35.8) | 219 (58.9) | 2,020 (44.1) | 949 (34.2) | 2,971 (30.0) |
| Beta-blocker | 6,017 (36.7) | 298 (80.1) | 2,132 (46.6) | 1,016 (56.0) | 2,755 (27.8) |
| TD | 4,002 (24.4) | 106 (28.5) | 1,318 (28.8) | 472 (26.0) | 2,223 (22.5) |
| Other antihypertensive<br>drug classes | 4,039 (24.7) | 227 (61.0) | 1,642 (35.9) | 936 (51.6) | 1,628 (16.4) |
| Combination anti-<br>hypertensive drug | 1,688 (10.3) | 40 (10.8) | 523 (11.4) | 138 (7.6) | 1,000 (10.1) |
| None | 4,886 (29.8) | 24 (6.5) | 905 (19.8) | 290 (16.0) | 3,657 (36.9) |

Abbreviations: ACEI, angiotensin-converting enzyme inhibitor; ARB, angiotensin-receptor blocker; BB, beta-blocker; CCB, calcium-channel blocker; IHD, ischemic heart disease; MI, myocardial infarction; CKD, chronic kidney disease; TD, thiazide or thiazide-like diuretic.
\* Uncomplicated hypertension was defined as lack of MI, coronary artery disease, diabetes, CKD, and cerebrovascular disease.

Among adults prescribed one drug class, ACEI or ARB was the most prescribed drug overall (35.9%) and among those with diabetes (46.3%), whereas BB (53.6%; 25.1%) were the most prescribed drugs for those with a history of MI and CKD, respectively (**Appendix Table 3**). Among adults prescribed two antihypertensive drug classes, most patients were prescribed an ACEI or ARB and a TD (21.0%), followed by ACEI or ARB and CCB (20.6%). Among patients using three or more antihypertensive drug classes, ACEI or ARB, TD, and CCB were most common (11.1%).

### Appropriate Pharmacological Treatment and Associated Factors

Based on the guideline-recommended antihypertensive drug classes by comorbidity described in **Appendix Table 2**, 67.0% of patients with prior MI, 51.4% of patients with diabetes, 48.2% of patients with CKD, and 49.2% of patients with uncomplicated hypertension were prescribed appropriate pharmacological treatment (**Figure 2**). In the subgroup analysis, the proportion of patients with appropriate pharmacological treatment was suboptimal (<70%) consistently across all demographic subgroups for patients who were post-MI, had diabetes, had CKD, or had uncomplicated hypertension (**Table 3**). No subgroup had an exceptionally high rate of appropriate pharmacological treatment.

**Table 3.** Proportion of patients with appropriate pharmacological treatment among treated patients by demographic subgroups.

|  | Comorbidity |  |  |  |  |
| --- | --- | --- | --- | --- | --- |
|  | All treated patients with any listed comorbidities (N= 10,324) | Prior MI (N=348) | Diabetes (N=3,672) | CKD (N=1,525) | Uncomplicated hypertension* (N=6,241) |
| **Age group, N (%)** |  |  |  |  |  |
| 18-44 years | 438 (50.3) | 7 (87.5) | 95 (54.0) | 27 (58.7) | 339 (49.2) |
| 45-64 years | 2,102 (52.3) | 71 (68.9) | 757 (54.3) | 201 (51.3) | 1,288 (50.7) |
| >=65 years | 2,670 (49.1) | 155 (65.4) | 1,035 (49.2) | 507 (46.6) | 1,444 (48.0) |
| **Sex, N (%)** |  |  |  |  |  |
| Female | 2,933 (51.3) | 124 (67.4) | 1,049 (53.9) | 384 (50.7) | 1,751 (49.0) |
| Male | 2,277 (49.4) | 109 (66.5) | 838 (48.5) | 351 (45.7) | 1,320 (49.5) |
| **Race, N (%)** |  |  |  |  |  |
| Black | 1,387 (56.7) | 45 (70.3) | 643 (59.2) | 240 (55.8) | 703 (54.4) |
| White | 3,136 (47.3) | 160 (66.1) | 903 (45.2) | 401 (43.3) | 2,040 (47.3) |
| Others | 627 (54.7) | 27 (67.5) | 332 (58.1) | 93 (57.8) | 278 (50.8) |
| Unknown | 60 (56.1) | 1 (50.0) | 9 (47.4) | 1 (14.3) | 50 (58.1) |
| **Ethnicity, N (%)** |  |  |  |  |  |
| Hispanic | 636 (56.0) | 33 (75.0) | 326 (57.4) | 84 (53.5) | 287 (53.1) |
| Non-Hispanic | 4,477 (49.8) | 198 (65.8) | 1,539 (50.3) | 647 (47.6) | 2,711 (48.9) |
| Other/Unknown | 97 (48.7) | 2 (66.7) | 22 (50.0) | 4 (44.4) | 73 (48.0) |
| **Insurance type, N (%)** |  |  |  |  |  |
| Public (Medicare or Medicaid) | 3,372 (50.5) | 178 (65.7) | 1357 (51.9) | 590 (48.1) | 1,818 (48.8) |
| Private | 1,644 (50.0) | 48 (70.6) | 446 (48.8) | 115 (46.4) | 1,153 (49.9) |
| Military | 8 (42.1) | NA (NA) | 4 (66.7) | 1 (100.0) | 3 (25.0) |
| None | 117 (56.8) | 6 (85.7) | 57 (65.5) | 16 (72.7) | 55 (47.8) |
| Others/Unknown | 69 (50.7) | 2 (50.0) | 23 (46.9) | 13 (46.4) | 42 (53.2) |
| **Preferred language, N (%)** |  |  |  |  |  |
| English | 4,741 (50.1) | 210 (67.3) | 1616 (50.3) | 657 (47.4) | 2,888 (49.2) |
| Spanish | 330 (56.8) | 17 (70.8) | 205 (60.3) | 54 (55.7) | 114 (50.0) |
| Others | 126 (49.8) | 6 (54.5) | 59 (52.7) | 22 (57.9) | 63 (48.5) |
| Unknown | 13 (50.0) | 0 (0.0) | 7 (70.0) | 2 (50.0) | 6 (42.9) |
| **BMI category, N (%)** |  |  |  |  |  |
| $\geq 30 \text{ kg/m}^2$ | 2,645 (53.1) | 99 (68.8) | 1131 (53.2) | 366 (53.0) | 1,414 (52.1) |
| 25-<30 $\text{kg/m}^2$ | 1,501 (49.0) | 77 (67.5) | 498 (50.3) | 208 (44.9) | 926 (47.8) |
| < 25 $\text{kg/m}^2$ | 907 (47.1) | 55 (64.0) | 212 (47.0) | 149 (44.7) | 621 (46.1) |
| Unknown | 157 (44.2) | 2 (50.0) | 46 (44.2) | 12 (31.6) | 110 (44.9) |
| **Smoking status, N (%)** |  |  |  |  |  |
| Current smoker | 243 (63.1) | 36 (72.0) | 97 (60.2) | 57 (64.0) | 57 (62.8) |
| Former smoker | 1043 (59.3) | 112 (77.2) | 513 (59.4) | 271 (57.4) | 194 (56.3) |
| Never smoker | 121 (53.8) | 19 (76.0) | 65 (54.6) | 29 (43.9) | 14 (48.2) |
| Unknown | 3803 (47.8) | 66 (51.6) | 1212 (47.9) | 378 (42.1) | 1912 (47.7) |

Abbreviations: MI, myocardial infarction; CKD, chronic kidney disease; BMI, body mass index.
\* Uncomplicated hypertension was defined as lack of MI, coronary artery disease, diabetes, CKD, and cerebrovascular disease.

**Figure 2.**
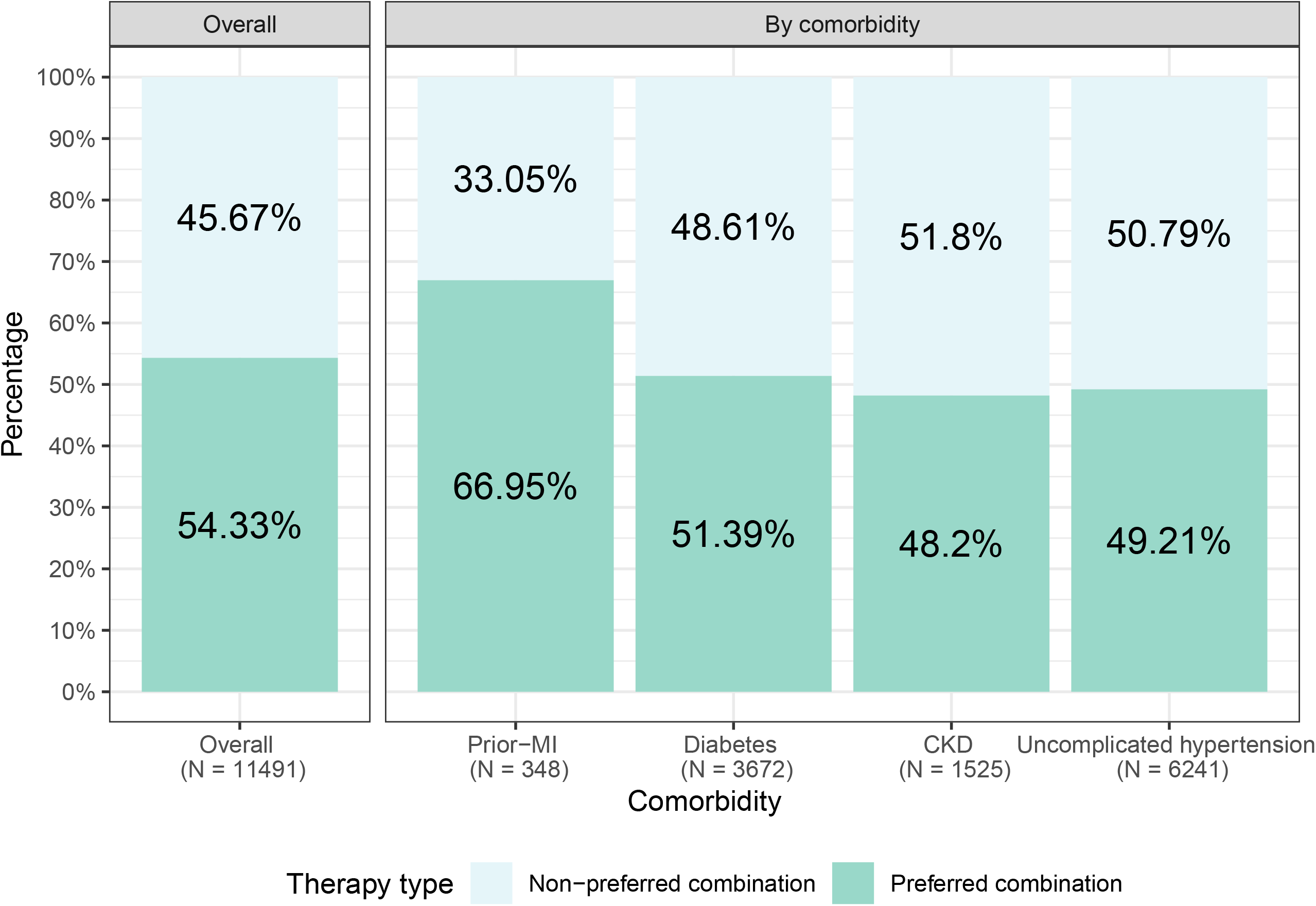
Distributions of patient-level and dyad-level measures of variation.

The gradient boosting machine learning model identified BMI, age, systolic and diastolic BP as more important factors associated with prescription of guideline-preferred combination therapy. Notably, prescription of guideline-preferred combination therapy was lower for patients with older age, lower BMI, and lower BP (**Figure 3**). Our sensitivity analysis using a different definition for active prescription showed results consistent with the main analysis (**Appendix Figures 5-6**).

**Figure 3.**
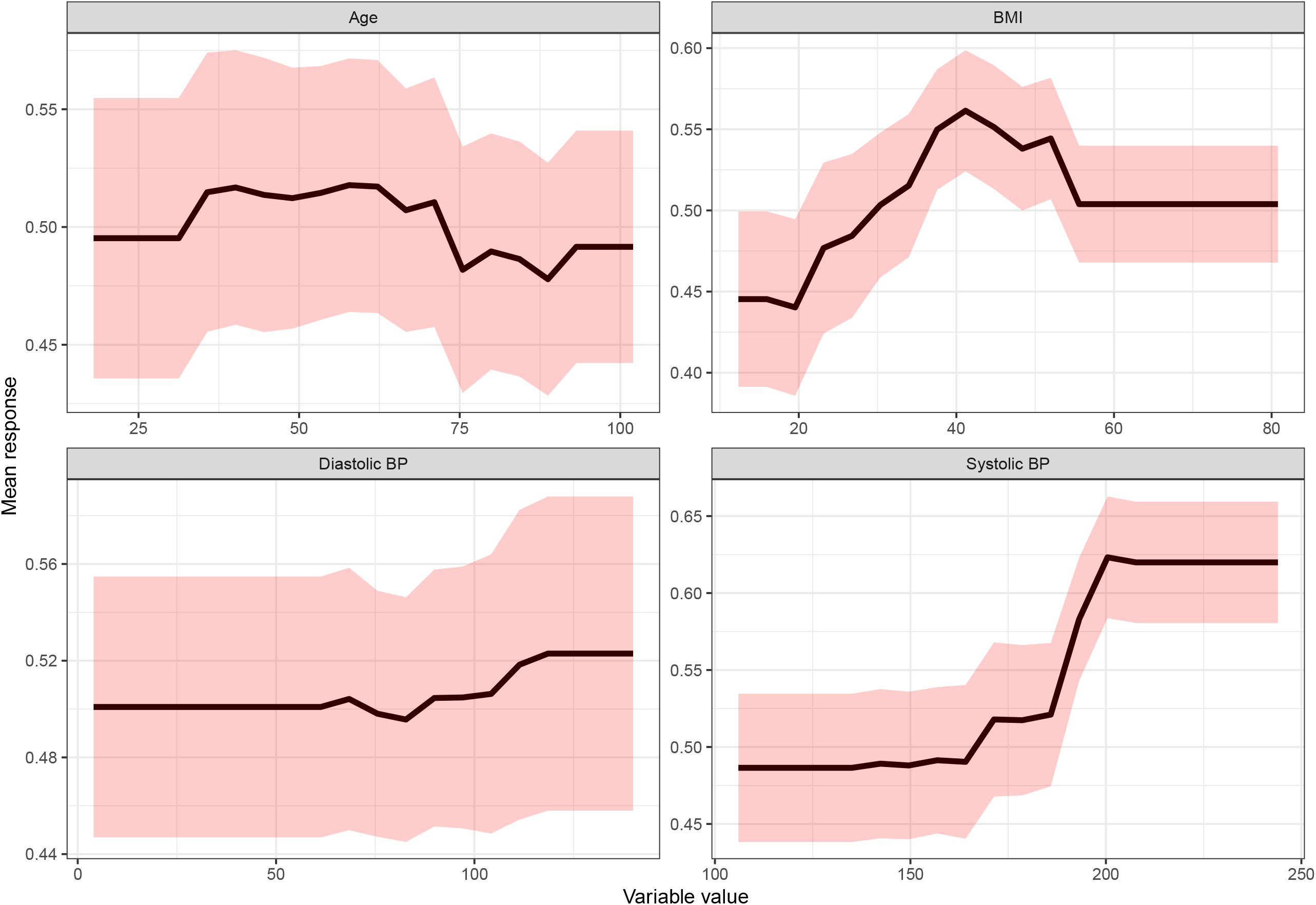
Partial dependency plots revealing association of prescribing preferred two drug class antihypertensive therapy with selected patient features. Abbreviations: BMI, body mass index; BP, blood pressure. Footnote: The partial dependence plots examined the average marginal effect that each feature has on the outcome variable. The x-axis showed the values of different feature variables (i.e., age, BMI, systolic blood pressure, diastolic blood pressure). The y-axis showed the probability of prescribing a guideline-recommended combination therapy given different values of the selected features.

## DISCUSSION

In this analysis of real-world data from a large health system, we found that about 30% of the patients with markedly elevated BP were not prescribed any antihypertensive drugs prior to 90 days of their second visit with BP ≥160/100 mmHg. Among those who were prescribed antihypertensive drugs, only half were prescribed guideline-recommended combination antihypertensive drugs. The suboptimal prescription of guideline-recommended combination therapy was consistent across age, gender, race, and other demographic subgroups. These findings indicate that major opportunities exist for improving the guideline adherence of antihypertensive drug prescription in this population.

This study extends the prior literature in two important ways. First, we used real-world clinical data to better characterize the prescription patterns of patients with markedly elevated BP in a large health system. Previous studies have shown that providers are not optimally titrating patient-specific combination therapy for hypertension.^6-9^ However, they focused on all hypertensive people or Medicare beneficiaries, were conducted more than a decade ago, or did not systematically assess medication intensity (number and class). Recent prescription data on those with markedly elevated BP are also limited. Our findings suggest that clinical inertia, defined as the provider’s failure to initiate or intensify therapy when the treatment goals are unmet,^17 18^ represent a significant barrier to hypertension control among patients with markedly elevated BP.

Second, we assessed the heterogeneity in prescription pattern across different patient subgroups defined by comorbidities and sociodemographic characteristics. We have shown that suboptimal guideline adherence to medication was common in high-risk patients with history of MI, diabetes, and CKD, in which the benefit of optimal treatment would be the greatest. These results are generally consistent with the prior studies that reported low guideline concordance in patients with comorbid risk factors,^8 9^ highlighting the need for effective approaches to eliminate clinical inertia in these patients. Of note, the proportion of patients treated with preferred two drug class antihypertensive therapy was higher among Black patients compared with White patients. This observation suggests that the racial disparity in BP control likely reflects other factors, such as differences in lifestyle and environmental stressors, access to or adherence with medications, or more treatment-resistant hypertension rather than clinical inertia.^19^

There may be multiple reasons why clinicians fail to intensify treatment (i.e., clinical inertia) for chronic conditions such as hypertension. First, the increasing demand on overloaded clinicians, the diffusion of responsibility, and short visit time may contribute to clinical inertia. In a recent survey of 1,273 healthcare professionals from 154 primary care practices, workplace burnout was reported in one third of the primary care physician and one fifth of the advanced practice clinicians.^20^ There is also diffusion of responsibility, where physicians tend to lessen their own sense of accountability by highlighting patient-level and system-level barriers.^21^ This issue is particularly pertinent given the time-constraints of clinical encounters. The average appointment time for a visit to a primary care physician is only 15 minutes. This short timeframe creates a challenge for physicians to weigh all the aspects of a patient’s condition and make the best treatment decisions. In addition, the lack of physician education and training of medical guideline may be another contributing factor to clinical inertia.^6^ For example, without effective education, physicians may not be aware of a lower treatment goal for patients with diabetes and certain medication classes recommended for African Americans. Many of the patients with markedly elevated BP may be seen by specialists who are not skilled in managing hypertension. Moreover, the large variation in office-based BP measurement and the uncertainty about patient’s true BP may also post a challenge in the treatment of hypertension. Previous studies have documented marked visit-to-visit variability in office-based BP values,^22-26^ which can overwhelm a signal of treatment response and impair clinical judgement. Other possible reasons include concerns about side effects and pessimism that control will be eventually achieved.^27^

Several strategies can be used on overcome clinical inertia in clinical practice. A systematic review of 10 randomized controlled trials aimed at reducing clinical inertia in the treatment of hypertension have shown that clinical decision support for intensification of treatment, ambulatory BP monitoring, and physician education interventions were associated with improvements in adherence to treatment guidelines and BP control.^28^ Additionally, implementing team-based care involving pharmacists, nurse practitioners, and community health workers in the diagnosis and management of hypertension is an effective strategy to improve BP control.^29 30^ Studies involving pharmacists to assist with titrating antihypertensive medication result in mean SBP reductions of 7.8 mm Hg (pharmacists in clinics) or 9.3 mm Hg (community pharmacists).^31^ Studies of community health workers, who serve as a link between the community and clinic, have also demonstrated effectiveness in managing hypertension and other chronic conditions.^32^ Furthermore, simplifying hypertension treatment algorithms can facilitate adherence to treatment guideline for hypertension.^3^ Practically, hypertension treatment protocols need to be implemented with consideration of local resources, challenges, and needs of patients and clinical care teams. Standardizing treatment algorithms to provide single targets for every level of hypertension and identifying easily obtained, objective metrics may boost the effectiveness of team strategies with clear goals. Finally, using performance and quality improvement metrics such as the 2019 AHA/ACC Clinical Performance and Quality Measures for Adults with High BP^33^ to evaluate provider performance may enhance accountability and reduce clinical inertia. This may include incorporating actionable decision-making information to identify people with uncontrolled hypertension into the EHR and designing next-generation quality improvement metrics to assess health system performance on hypertension control.^34^

## Limitations

This study has several limitations. First, these data are not necessarily generalizable to other health systems. Second, only one federally qualified health center uses the YNHHS EHR; thus, we are likely missing some patients with markedly elevated BP who are being treated in the community. However, as YNHHS is the largest health system in Connecticut, the vast majority of treatment within this geographic area occurs within YNHHS. Third, we used data from only outpatient settings and excluded potential patients with markedly elevated BP in which care was received only in an inpatient or emergency department setting; however, BP elevations in these acute settings are less likely to be indicative of the patients’ true BP. Finally, treatment of hypertension was ascertained based on medication prescription data and we did not have data on patient adherence to medication.

## Conclusion

In conclusion, use of hypertension guidelines is suboptimal and a substantial proportion of adults with markedly elevated BP receive no or only one class of antihypertensive medications, even though clinicians can titrate these treatments or add a second medication to improve hypertension control. Major opportunities exist for improving the guideline adherence of antihypertensive drug prescription in this population.

## Data Availability

Technical appendix, statistical code, and dataset available from the corresponding author.

## Competing interests

All authors have completed the ICMJE uniform disclosure form at www.icmje.org/coi_disclosure.pdf and declare: Dr. Lu is supported by the National Heart, Lung, and Blood Institute (K12HL138037) and the Yale Center for Implementation Science. In the past three years, Dr. Krumholz received expenses and/or personal fees from UnitedHealth, Element Science, Aetna, Reality Labs, Johnson & Johnson, the Siegfried and Jensen Law Firm, Arnold and Porter Law Firm, Martin/Baughman Law Firm, and F-Prime. He is a co-founder of Refactor Health and HugoHealth, and is associated with contracts, through Yale New Haven Hospital, from the Centers for Medicare & Medicaid Services. Dr. Schulz collaborates with the National Center for Cardiovascular Diseases in Beijing; is a technical consultant to HugoHealth, a personal health information platform, and co-founder of Refactor Health, an AI-augmented data management platform for healthcare; is a consultant for Interpace Diagnostics Group, a molecular diagnostics company. Dr. Ross currently receives research support through Yale University from Johnson and Johnson to develop methods of clinical trial data sharing, from the Medical Device Innovation Consortium as part of the National Evaluation System for Health Technology (NEST), from the Food and Drug Administration for the Yale-Mayo Clinic Center for Excellence in Regulatory Science and Innovation (CERSI) program (U01FD005938); from the Agency for Healthcare Research and Quality (R01HS022882), from the National Heart, Lung and Blood Institute of the National Institutes of Health (NIH) (R01HS025164, R01HL144644), and from the Laura and John Arnold Foundation to establish the Good Pharma Scorecard at Bioethics International. In the past 36 months, NDS has received research support through Mayo Clinic from the Food and Drug Administration to establish Yale-Mayo Clinic Center for Excellence in Regulatory Science and Innovation (CERSI) program (U01FD005938); the Centers of Medicare and Medicaid Innovation under the Transforming Clinical Practice Initiative (TCPI); the Agency for Healthcare Research and Quality (R01HS025164; R01HS025402; R03HS025517; K12HS026379); the National Heart, Lung and Blood Institute of the National Institutes of Health (NIH) (R56HL130496; R01HL131535; R01HL151662); the National Science Foundation; from the Medical Device Innovation Consortium as part of the National Evaluation System for Health Technology (NEST)and the Patient Centered Outcomes Research Institute (PCORI) to develop a Clinical Data Research Network (LHSNet).

## Copyright

The Corresponding Author has the right to grant on behalf of all authors and does grant on behalf of all authors, an exclusive license on a worldwide basis to the BMJ Publishing Group Ltd to permit this article (if accepted) to be published in BMJ editions and any other BMJPGL products and sublicenses such use and exploit all subsidiary rights.

## Details of contributors

All authors are responsible for study concept and design. YL, CH, YLiu were responsible for the acquisition of data and its analysis. All Authors contributed to the interpretation of the data. YL drafted the manuscript. All authors performed critical revision of the manuscript for important intellectual content and approved the final manuscript. HMK is the guarantor.

## Funding

This research received no specific grant from any funding agency in the public, commercial, or not-for-profit sectors.

## Ethical approval

Not required.

## Data sharing

Technical appendix, statistical code, and dataset available from the corresponding author.

## Transparency

The lead author affirms that this manuscript is an honest, accurate, and transparent account of the study being reported; that no important aspects of the study have been omitted; and that any discrepancies from the study as planned (and, if relevant, registered) have been explained.

## Patient involvement

No patients were asked for input in the creation of this article.

## Notes

### Funding Statement

This study did not receive any funding

### Author Declarations

This study received an exemption for review from the Institutional Review Board at Yale University because EHR data are de-identified.

