## Supplemental material for "Medication Guideline Adherence Among Patients with Markedly Elevated Blood Pressure in A Real-World Setting"

**Online Supplement**

**Table of Contents**

| **Content** | **First Page** |
| --- | --- |
| **Appendix Figure 1.** Schema of target cohort definition. | 2 |
| **Appendix Figure 2.** Two approaches for defining active antihypertensive prescription. | 3 |
| **Appendix Figure 3.** Performance of different approaches identifying patients with antihypertensive prescription. | 4 |
| **Appendix Figure 4.** Flow diagram showing selection of study population. | 5 |
| **Appendix Figure 5.** Number of antihypertensive medication class prescribed among patients with markedly elevated blood pressure in the sensitivity analysis. | 6 |
| **Appendix Figure 6.** Proportion of patients treated with preferred vs. non-preferred two drug class antihypertensive therapy among treated patients in the sensitivity analysis. | 7 |
| **Appendix Table 1.** Diagnosis codes for comorbidities. | 8 |
| **Appendix Table 2.** Appropriate pharmacological treatment for patients with specific comorbidity. | 9 |
| **Appendix Table 3.** Top three commonly prescribed antihypertensive medication classes among patients with markedly elevated blood pressure. | 10 |
| **Appendix Table 4.** Antihypertensive medication classes prescribed among patients with markedly elevated blood pressure in sensitivity analysis. | 12 |

**Appendix Figure 1.** Schema of target cohort definition.

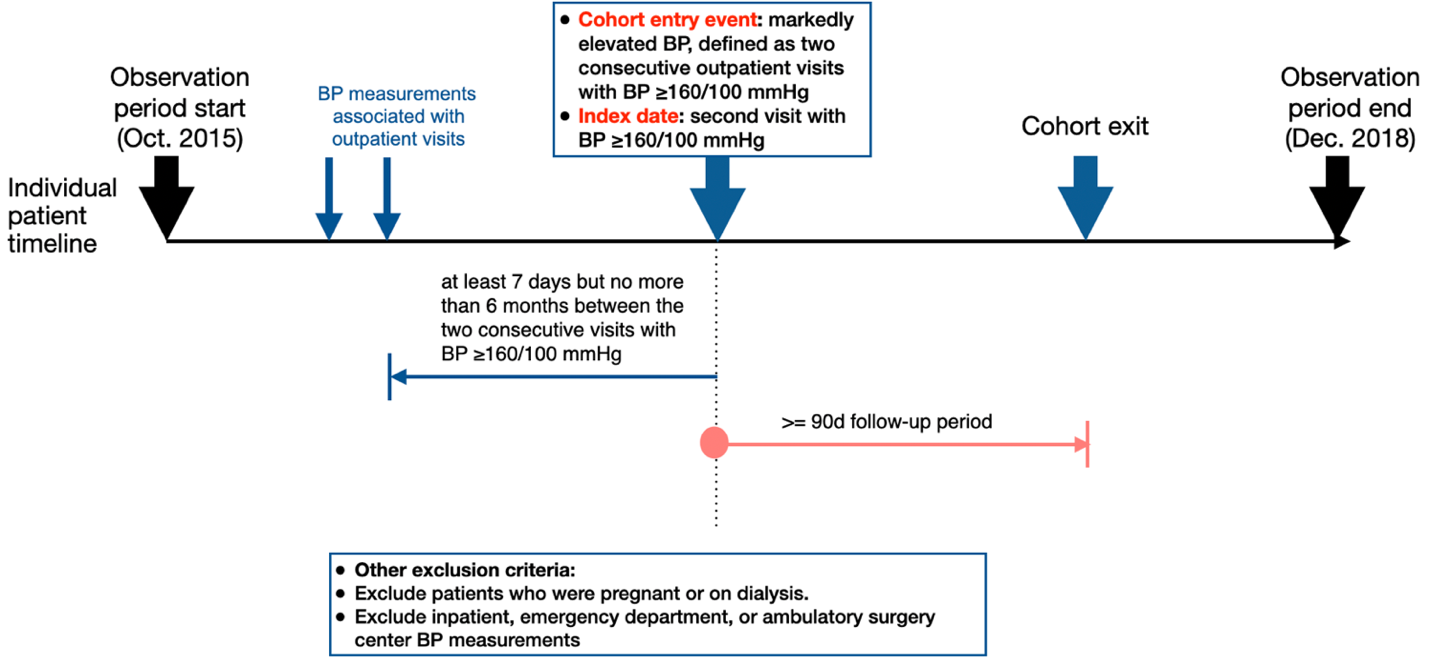

**Appendix Figure 2.** Two approaches for defining active antihypertensive prescription.

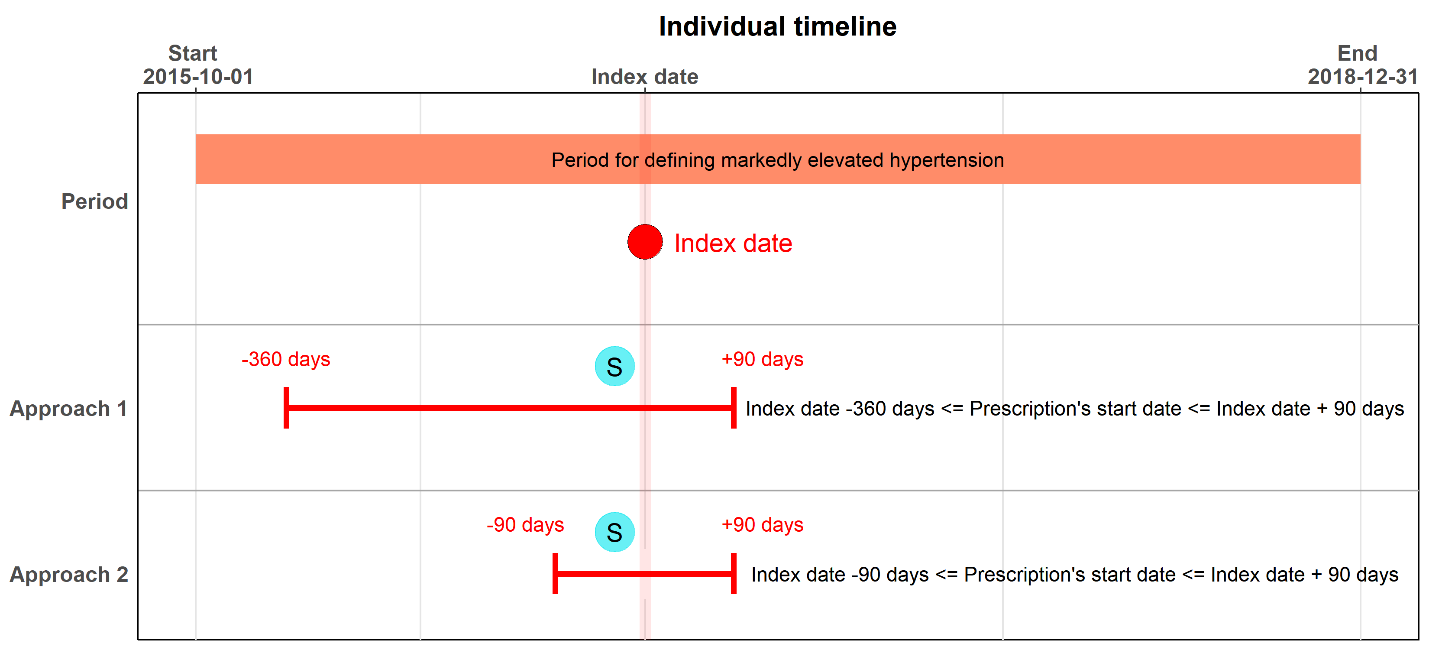

**Appendix Figure 3.** Performance of different approaches identifying patients with antihypertensive prescription.

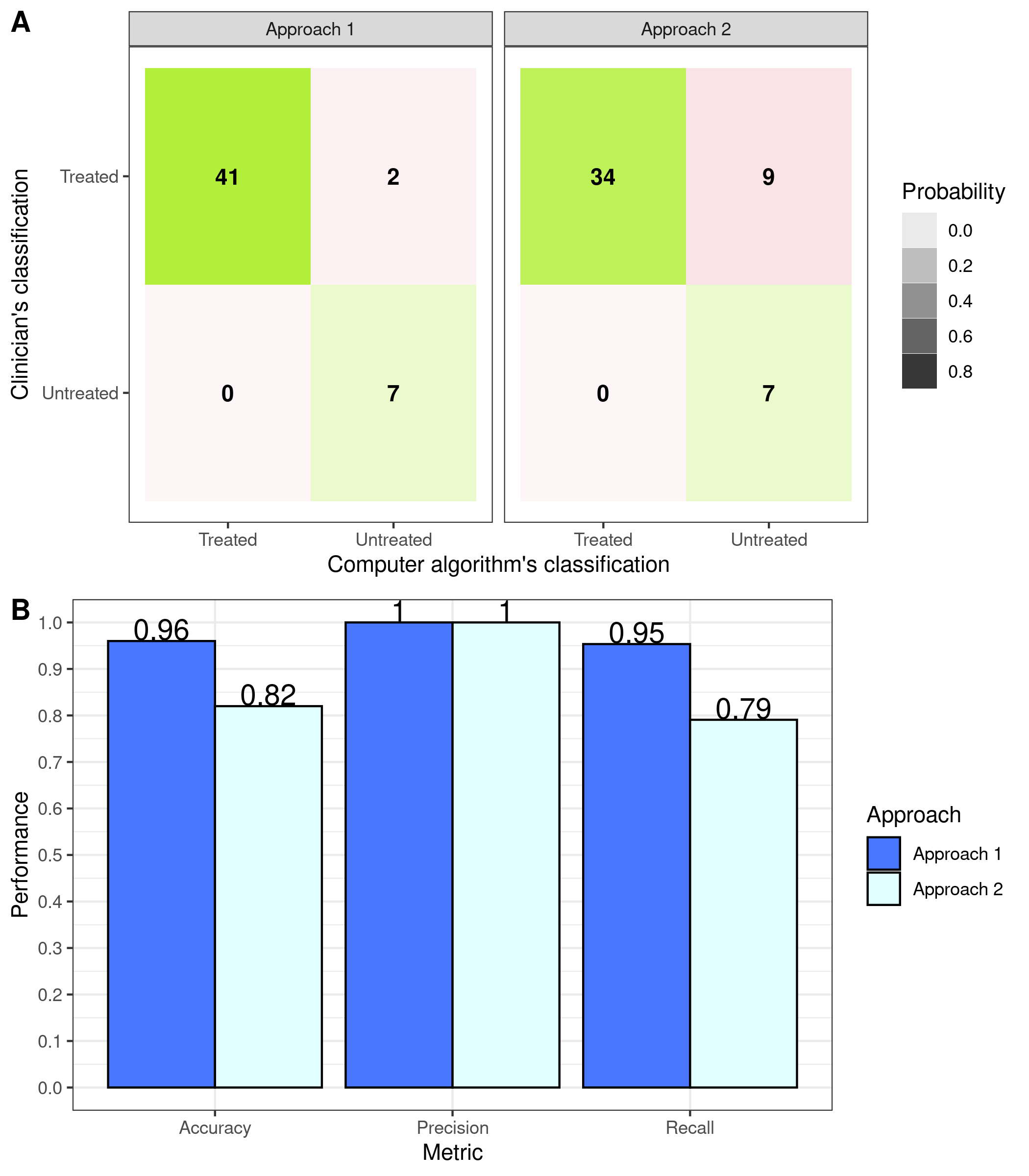

**Appendix Figure 4.** Flow diagram showing selection of study population.

Unique patients aged >=18 years who had at least two outpatient visits in YNHHS from October 1^st^, 2015 to December 31^st^, 2018

N = 627,361

Patients aged >=18 years who had at least two consecutive outpatient visits with BP >= 160/100 mmHg

N =28,590

Patients who did not have at least two consecutive outpatient visits with BP>=160/100 mmHg

N = 598,771

Patients who had less than 7 days or more than 6 months between the two consecutive visits with BP>=160/100 mmHg

N = 6,267

Patients aged >=18 years who had at least two consecutive outpatient visits (at least 7 days but no more than 6 months apart) with BP >= 160/100 mmHg

N =22,323

Patients aged 18-85 years who had at least two consecutive outpatient visits (at least 7 days but no more than 6 months apart) with BP >= 160/100 mmHg and 3-month follow-up time (after Oct 1^st^, 2016)

N =17,415

Patients who had less than 3 months follow-up

N = 4,908

Patients included in the analysis

N =16,377

Patients with pregnancy or on dialysis

N = 1,038

**Appendix Figure 5.** Number of antihypertensive medication class prescribed among patients with markedly elevated blood pressure in the sensitivity analysis.

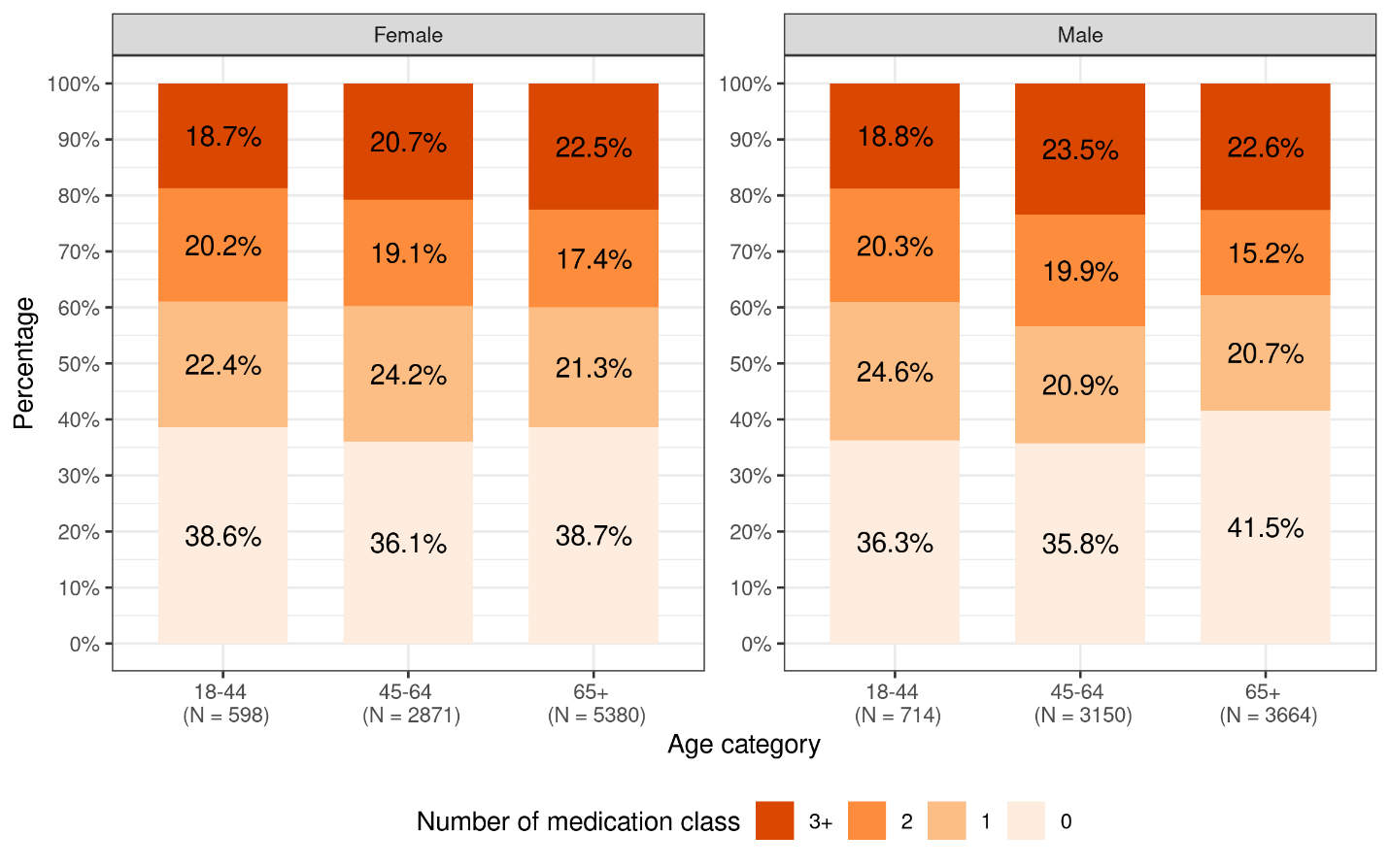

Footnote: In the sensitivity analysis, active prescription was defined when the prescription order start date was between 90 days prior to the index date and 90 days after the index date.

**Appendix Figure 6.** Proportion of patients treated with preferred vs. non-preferred two drug class antihypertensive therapy among treated patients in the sensitivity analysis.

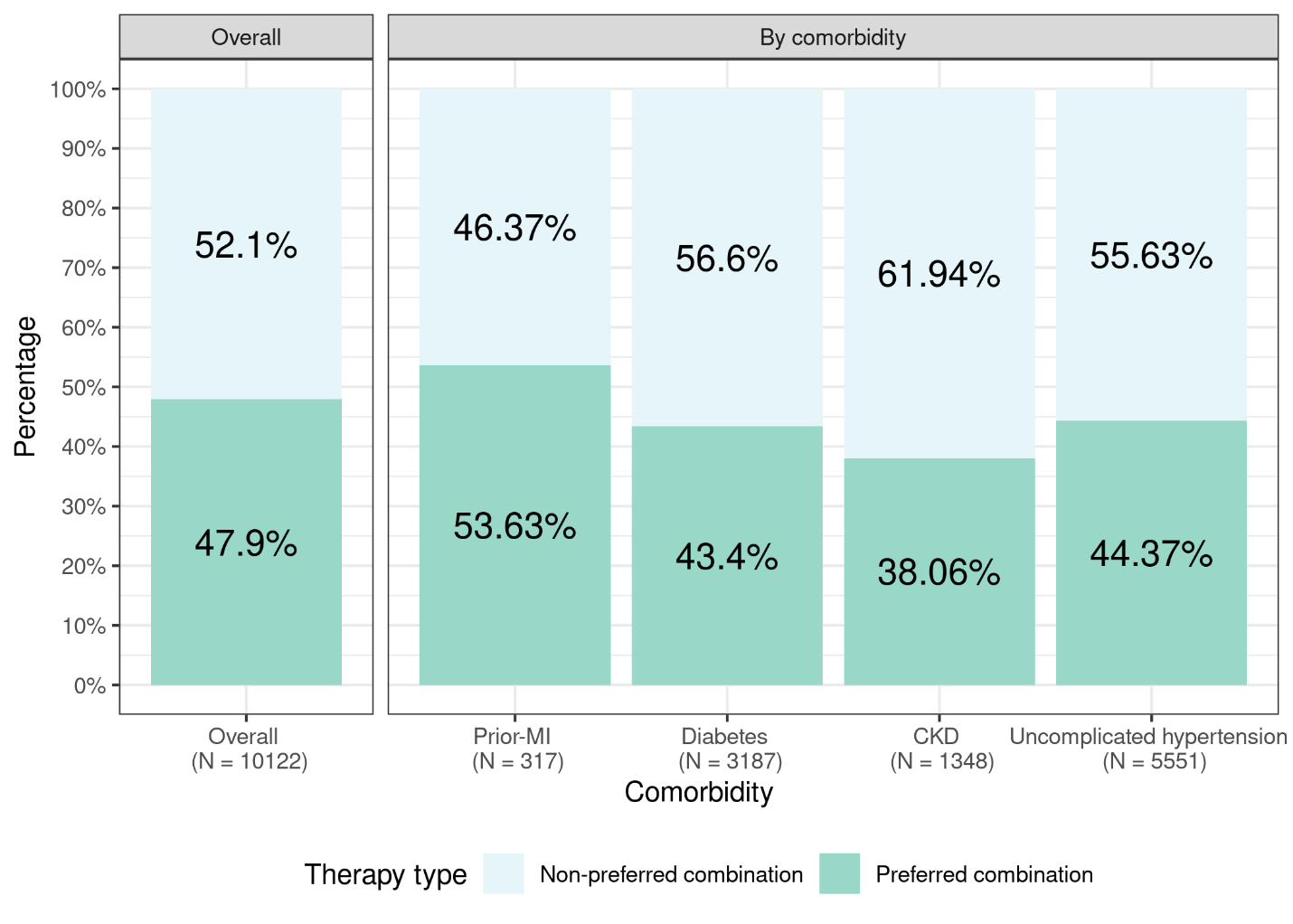

Footnote: In the sensitivity analysis, active prescription was defined when the prescription order start date was between 90 days prior to the index date and 90 days after the index date.

**Appendix Table 1.** Diagnosis codes for comorbidities.

| **Comorbidity** | **ICD-10-CM code** |
| --- | --- |
| Heart failure | I09.9 I11.0 I13.0 I13.2 I25.5 I42.0 I42.5 I42.6 I42.7 I42.8 I42.9 I43 I50 |
| Diabetes mellitus | E10 E11 E12 E13 E14 E10.69 E11.69 |
| Dyslipidemia | E78.5 E78.2 E78.4 |
| Acute myocardial infarction | I21 I22 |
| Coronary artery disease | I25.1 |
| Cerebrovascular disease | I67.89 I67.9 I67.858 I69 |
| Atrial fibrillation/Atrial Flutter | I48 |
| Chronic kidney disease | N18 I12 I13 |
| Chronic obstructive pulmonary disease | J44 |
| Peripheral arterial disease | I73 |
| Angina | I20 I20.0 I20.1 I20.8 I20.9 I23.7 |
| Hemorrhagic stroke | I60 I61 I69.0 I69.1 |
| Ischemic stroke | I63 I69.3 |
| Depression | F32 F32.3 F32.8 F32.89 F32.9 F33 F33.3 F33.4 |
| Dementia | F01 F01.5 F02.8 F03 F03.9 |
| Hypertensive retinopathy | H35.03 |
| Substance use disorder | F10, F11, F12, F13, F14, F15, F16, F17, F18, F19 |

**Appendix Table 2.** Appropriate pharmacological treatment for patients with specific comorbidity.

| Comorbidity | Preferred drug class | Non-preferred drug class |
| --- | --- | --- |
| Prior MI | ACEI/ARB + BB | All other combinations |
| Diabetes | ACEI/ARB + CCB/TD | All other combinations |
| CKD | ACEI/ARB + CCB/TD | All other combinations |
| Uncomplicated hypertension | ACEI/ARB + CCB/TD or BB + CCB/TD | All other combinations |

ACEI/ARB refers to either ACEI or ARB; CCB/TD refers to either CCB or TD.

Abbreviations: ACEI, angiotensin-converting enzyme inhibitor; ARB, angiotensin-receptor blocker; BB, beta-blocker; CCB, calcium-channel blocker; IHD, ischemic heart disease; MI, myocardial infarction; CKD, chronic kidney disease; TD, thiazide or thiazide-like diuretic.

**Appendix Table 3.** Top three commonly prescribed antihypertensive medication classes among patients with markedly elevated blood pressure.

| Medication class | Comorbidity | | | | |
| --- | --- | --- | --- | --- | --- |
|  | All with marked elevated BP | Post-MI | Diabetes | CKD | Uncomplicated hypertension |
| Among adults using one medication class | | | | | |
| 1^st^ | ACEI or ARB | BB | ACEI or ARB | BB | ACEI or ARB |
|  | 1,176 (35.9) | 15 (53.6) | 353 (46.3) | 63 (25.1) | 739 (33.7) |
| 2^nd^ | BB | ACEI or ARB | BB | ACEI or ARB | CCB |
|  | 796 (24.3) | 7 (25.0) | 154 (20.2) | 63 (25.1) | 545 (24.8) |
| 3^rd^ | CCB | CCB | CCB | CCB | BB |
|  | 742 (22.6) | 4 (14.3) | 130 (17.1) | 55 (21.9) | 515 (243.5) |
| Total | 3,277 | 28 | 762 | 251 | 2,194 |
| Among adults using two medication class | | | | | |
| 1^st^ | ACEI or ARB and TD | ACEI or ARB and BB | ACEI or ARB and CCB | ACEI or ARB and CCB | ACEI or ARB and TD |
|  | 673 (21.0) | 17 (29.3) | 198 (21.6) | 57 (18.9) | 468 (24.3) |
| 2^nd^ | ACEI or ARB and CCB | CCB and BB | ACEI or ARB and BB | CCB and BB | ACEI or ARB and CCB |
|  | 658 (20.6) | 12 (20.7) | 174 (19.0) | 46 (15.3) | 404 (20.9) |
| 3^rd^ | ACEI or ARB and BB | Other diuretics and BB | ACEI or ARB and TD | ACEI or ARB and BB | ACEI or ARB and BB |
|  | 531 (16.6) | 8 (13.8) | 166 (18.1) | 42 (14.0) | 268 (13.9) |
| Total | 3,199 | 58 | 915 | 301 | 1,929 |
| Among adults using three or more medication classes | | | | | |
| 1^st^ | ACEI or ARB, TD, CCB | ACEI or ARB, BB, CCB | ACEI or ARB, TD, CCB | ACEI or ARB, BB, CCB | ACEI or ARB, TD, CCB |
|  | 555 (11.1) | 25 (9.5) | 195 (9.8) | 74 (7.6) | 327 (15.4) |
| 2^nd^ | ACEI or ARB, BB, CCB | ACEI or ARB, Other diuretic, BB, CCB | ACEI or ARB, BB, CCB | ACEI or ARB, Other diuretic, BB, CCB | ACEI or ARB, BB, CCB |
|  | 514 (10.2) | 21 (8.0) | 181 (9.1) | 66 (6.8) | 235 (11.1) |
| 3^rd^ | ACEI or ARB, TD, BB, CCB | ACEI or ARB, Other diuretics, BB, CCB, Arteriolar Smooth muscle agents | ACEI or ARB, TD, BB, CCB | ACEI or ARB, Other diuretics, BB, CCB, Arteriolar Smooth muscle agents | ACEI or ARB, TD, BB, CCB |
|  | 410 (8.2) | 20 (7.6) | 153 (7.7) | 43 (4.4) | 195 (9.2) |
| Total | 5,015 | 262 | 1,995 | 973 | 2,118 |

Abbreviations: BP, blood pressure; ACEI, angiotensin-converting enzyme inhibitor; ARB, angiotensin-receptor blocker; BB, beta-blocker; CCB, calcium-channel blocker; TD, thiazide or thiazide-like diuretic.

* Uncomplicated hypertension was defined as lack of MI, coronary artery disease, diabetes, CKD, and cerebrovascular disease. The four comorbidity groups are not mutually exclusive. It is possible that a patient has more than one condition.

**Appendix Table 4.** Antihypertensive medication classes prescribed among patients with markedly elevated blood pressure in sensitivity analysis.

| Medication class | All with marked elevated BP | Post-MI | Diabetes | CKD | Uncomplicated hypertension |
| --- | --- | --- | --- | --- | --- |
| ACEI | 3,337 (20.4) | 127 (34.1) | 1,213 (26.5) | 387 (21.3) | 1,696 (17.1) |
| ARB | 2,928 (17.9) | 107 (28.8) | 966 (21.1) | 380 (20.9) | 1,526 (15.4) |
| ACEI or ARB | 6,006 (36.7) | 222 (59.7) | 2,096 (45.8) | 737 (40.6) | 3,085 (31.2) |
| CCB | 4,790 (29.2) | 170 (45.7) | 1,603 (35.0) | 756 (41.7) | 2,483 (25.1) |
| Beta-blocker | 4,729 (28.9) | 235 (63.2) | 1,673 (36.6) | 799 (44.0) | 2,188 (22.1) |
| TD | 3,200 (19.5) | 76 (20.4) | 979 (21.4) | 326 (18.0) | 1,867 (18.9) |
| Other antihypertensive drug classes | 3,130 (19.1) | 175 (47.0) | 1,277 (27.9) | 735 (40.5) | 1,272 (12.9) |
| Combination anti-hypertensive drug | 1,338 (8.2) | 30 (8.1) | 383 (8.4) | 96 (5.3) | 829 (8.4) |
| None | 6,255 (38.2) | 55 (14.8) | 1,390 (30.4) | 467 (25.7) | 4,347 (43.9) |

Abbreviations: BP, blood pressure; ACEI, angiotensin-converting enzyme inhibitor; ARB, angiotensin-receptor blocker; BB, beta-blocker; CCB, calcium-channel blocker; IHD, ischemic heart disease; MI, myocardial infarction; CKD, chronic kidney disease; TD, thiazide or thiazide-like diuretic.

* Uncomplicated hypertension was defined as lack of MI, coronary artery disease, diabetes, CKD, and cerebrovascular disease. The four comorbidity groups are not mutually exclusive. It is possible that a patient has more than one condition.
